# Anaesthesia and delirium: a Mendelian randomization study

**DOI:** 10.1101/2024.06.02.24308334

**Authors:** YanHui Li, Yingwei Sun, Chuanyang Zhou, Lei Tan

## Abstract

**Background:** Delirium is a frequent complication in hospitalized older adults post-surgery associated with adverse outcomes. Although anaesthesia is traditionally linked to increased delirium risk, the causal relationship remains uncertain.

**Methods:** We conducted Mendelian randomization (MR) analyses using genome-wide association studies (GWAS) summary statistics to explore the causal effects of different anaesthesia types (general, regional, and local) on delirium risk. We employed the weighted median, MR-Egger, and MR-PRESSO methods for estimation and conducted sensitivity analyses to address pleiotropy and heterogeneity.

**Results:** Genetically determined anaesthesia types showed no significant causal effect on delirium risk. Sensitivity analyses confirmed the robustness of these findings, with no evidence of horizontal pleiotropy or significant heterogeneity.

**Conclusions:** Mendelian randomization provides strong evidence against a causal link between genetically determined anaesthesia and increased delirium risk.

## Introduction

Delirium, commonly seen in hospitalized older adults’ post-acute illness or surgery, involves sudden attention and cognitive function decline^1,2^ Its incidence ranges from 11% to 51%, varying by clinical setting and patient demographics^3,4^. Delirium is associated with adverse outcomes like extended hospital stays, higher morbidity and mortality, and cognitive decline^5^. It often arises from underlying conditions like diseases, metabolic disorders, and medication usage^1^. Despite its clinical significance, the exact mechanisms and effective interventions remain unclear^1,6^. Timely recognition and intervention are crucial, given delirium’s early onset post-surgery and varying incidence rates^7^. Addressing delirium promptly is vital to prevent long-term cognitive impairment and other serious health issues.

Anaesthesia, a crucial aspect of modern medicine, induces temporary loss of sensation or consciousness for pain-free medical procedures. Since the introduction of ether anaesthesia in 1846, it has become indispensable for surgeries, ranging from minor to complex procedures like organ transplants^8,9^. Techniques include general, regional and local anaesthesia, selected based on procedure type, patient health, and preferences. Challenges persist in managing complex cases and ensuring safety, with risks such as adverse reactions to anaesthetic agents being concerns^10^. Clinical research suggests a significant link between anaesthesia and delirium^11,12^. However, due to confounding factors such as age, anaemia, medication, and others, the causal relationship between anaesthesia and delirium remains unclear^13^. Moreover, limited studies have explored the impacts of different anaesthetic methods on delirium risk.

Mendelian Randomization (MR) studies use genetic variants as instrumental variables to evaluate causal relationships between exposures and outcomes, similar to randomized controlled trials in observational data. By utilizing the random assortment of genetic variants during meiosis, MR offers strong evidence for causal inference, addressing concerns like reverse causation and confounding^14^. This approach provides insights into the potential effects of interventions or exposures on outcomes, facilitating the identification of therapeutic targets and informing public health policies^15^. MR is particularly beneficial for investigating causal links between anaesthesia and delirium, serving as an efficient alternative to randomized controlled trials.

In this study, we utilized Two-Sample Mendelian randomization (TSMR) analyses to evaluate the impact of different forms of anaesthesia, including general, regional, and local anaesthesia, on the risk of delirium.

## Methods

### 1. Study design and data sources

In this study, we employed MR analyses to investigate the causal effects of different forms of anaesthesia on delirium risk. Adhering to three core assumptions ensured the validity of our results: (1) establishing a reliable association between genetic variants and the risk factor; (2) confirming no association between genetic variants and confounders; and (3) ensuring genetic variants solely influence the outcome through the risk factors^16^. A study design flowchart is provided in Figure 1. Ethical approval was not required as publicly available data were used, and reporting followed the STROBE-MR guidelines^17^.

**Figure 1:**
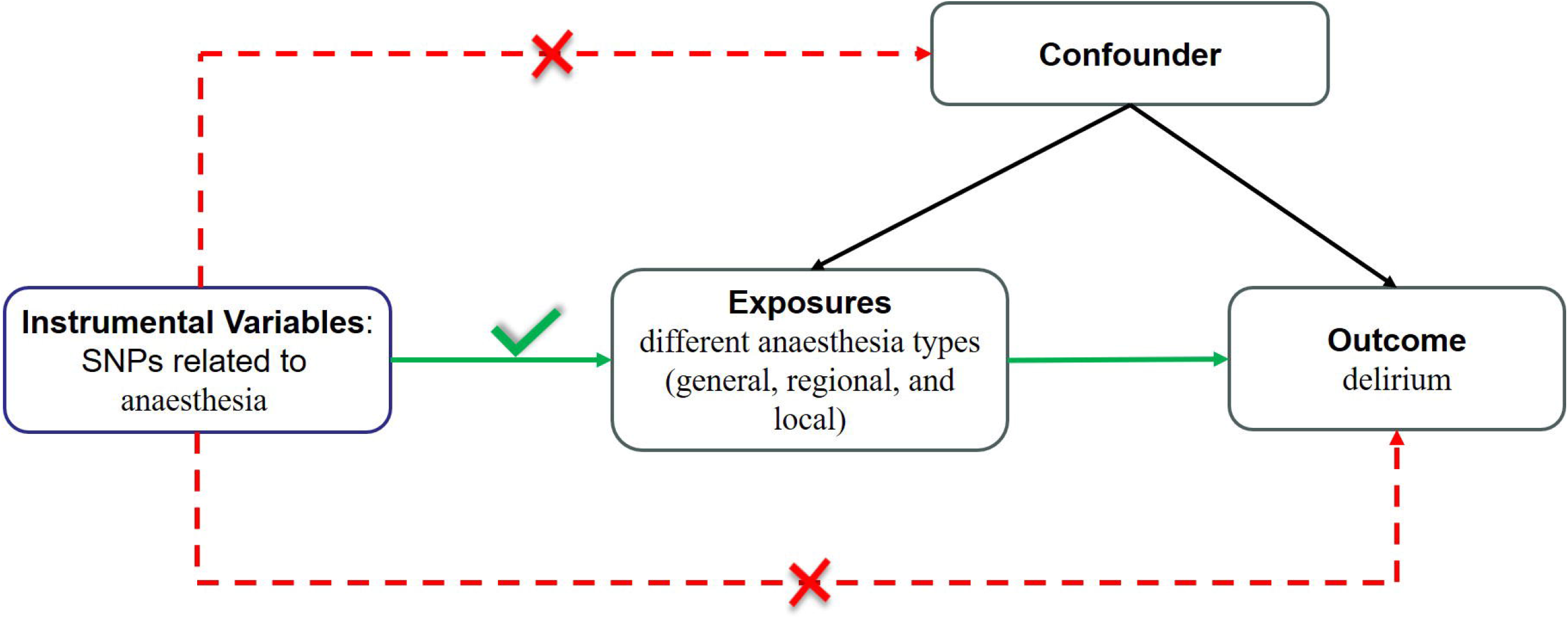
Study design flowchart of the Mendelian Randomization study. GWAS, genome-wide association studies; Anaes.:Anaesthesia, ; IVW: Inverse variance weighted; IVs: Instrumental variables; SNPs: Single Nucleotide Polymorphisms; MR, Mendelian randomization; TSMR, Two-Sample Mendelian randomization; OR: Odds ratio; CI, confidence interval; GWAS: Genome-Wide Association Study; UKBB, United Kingdom Biobank.

Our study utilized the most recent and comprehensive publicly available summary statistics from multiple genome-wide association studies (GWAS) sources, including the FinnGen database, the UK Biobank (UKBB) database. To avoid overlap, we specifically selected anaesthesia data from UKBB and delirium data from FinnGen as the subjects of our research. Detailed information on the GWAS data used in this study is provided in Supplementary Table 1.

**Table 1:**
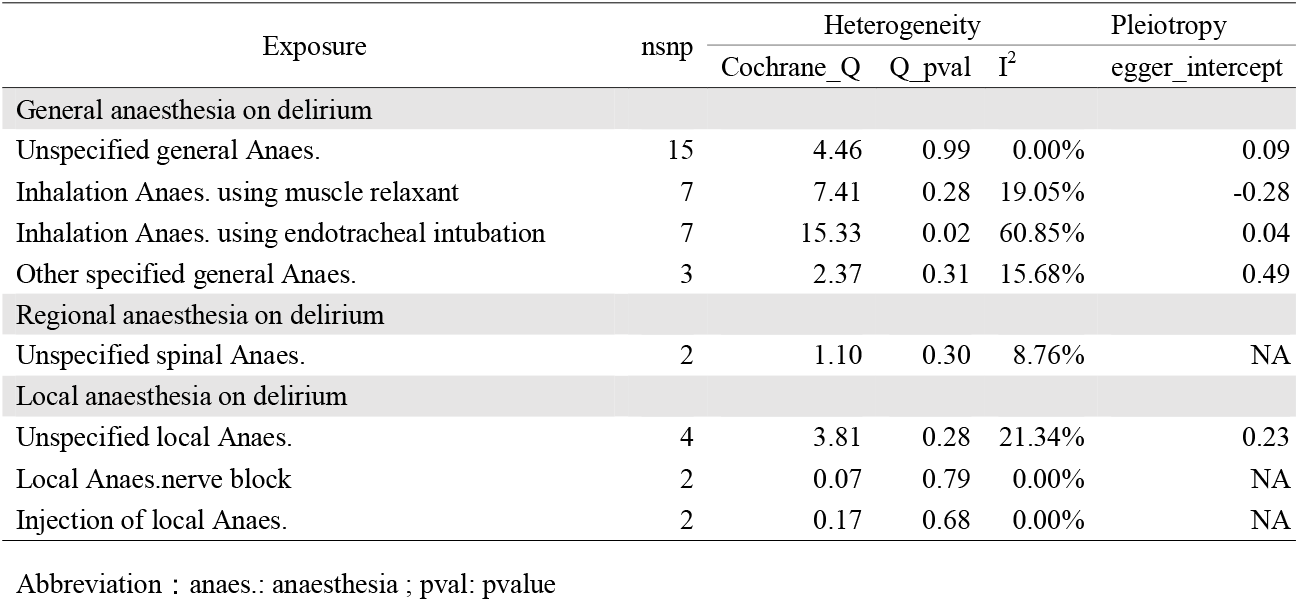
Sensitivity analysis of anaesthesia with the risk of delirium.

| Exposure | nsnp | Heterogeneity |  |  | Pleiotropy |
| --- | --- | --- | --- | --- | --- |
|  |  | Cochrane_Q | Q_pval | I<sup>2</sup> | egger_intercept |
| General anaesthesia on delirium |  |  |  |  |  |
| Unspecified general Anaes. | 15 | 4.46 | 0.99 | 0.00% | 0.09 |
| Inhalation Anaes. using muscle relaxant | 7 | 7.41 | 0.28 | 19.05% | -0.28 |
| Inhalation Anaes. using endotracheal intubation | 7 | 15.33 | 0.02 | 60.85% | 0.04 |
| Other specified general Anaes. | 3 | 2.37 | 0.31 | 15.68% | 0.49 |
| Regional anaesthesia on delirium |  |  |  |  |  |
| Unspecified spinal Anaes. | 2 | 1.10 | 0.30 | 8.76% | NA |
| Local anaesthesia on delirium |  |  |  |  |  |
| Unspecified local Anaes. | 4 | 3.81 | 0.28 | 21.34% | 0.23 |
| Local Anaes.nerve block | 2 | 0.07 | 0.79 | 0.00% | NA |
| Injection of local Anaes. | 2 | 0.17 | 0.68 | 0.00% | NA |

Abbreviation : anaes.: anaesthesia ; pval: pvalue

### 2. Genetic association datasets

#### GWAS data for anaesthesia

The summary data for the GWAS were obtained from the Medical Research Council-integrative Epidemiology Unit (MRC-IEU) using data from the UKBB. Anaesthesia techniques were categorized based on the Office of Population Censuses and Surveys Classification of Interventions and Procedures (OPCS-4), which includes general anaesthesia, regional anaesthesia, and local anaesthesia. Detailed information about the single nucleotide polymorphisms (SNPs) utilized in this study can be found in Supplementary Tables 3, 5 and 7.

#### GWAS data for Outcomes

The study investigated delirium as the outcome. Genetic data were sourced from the FinnGen consortium as of June 2021, involving 1269 cases and 209,487 controls of Finnish ancestry, identifying approximately 16 million SNPs. Delirium diagnoses were based on ICD-10 codes, including factors such as dementia combined with delirium, postoperative delirium, and other unspecified types, but excluding alcohol and psychoactive drug-induced delirium. Detailed information is available at https://r7.risteys.finngen.fi/.

### 3. Genetic Instrument Selection

For genetic instrument selection, due to the limited number of SNPs identified for anaesthesia as the exposure, a higher significance threshold (*p* < 1e-6) was applied. Variants meeting this threshold were clumped for linkage disequilibrium (LD) using a distance window of 10,000 kB and an r^2^ <0.001. To prevent weak instrumental bias, the F statistic was calculated to evaluate the strength of the instrumental variables (IV). An F > 10 indicated a robust association between the IV and exposures, thus safeguarding the MR analysis against potential weak instrumental bias. After several rounds of rigorous filtering, a set of eligible instrumental variables for the subsequent MR analysis was obtained.

### 4. Statistical analysis

We utilized the “TwoSampleMR” ^16^, “ MendelianRandomization “ ^18^, and “MR-PRESSO”^16^ packages for MR analyses, incorporating sensitivity tests. Causal estimates were reported as odds ratios (ORs) with 95% confidence intervals (CIs). Statistical analyses were performed using R software version 4.3.2 (The R Foundation for Statistical Computing).

#### Two-Sample MR analysis

Causal effects were estimated using the random-effects inverse variance weighted (IVW) method ^19^. To ensure unbiased estimates, MR analyses were also conducted using four alternative methods (MR Egger, Simple mode, Weighted median, and Weighted mode) when feasible. A causal effect was considered suggested if the IVW p-value was less than 0.05. Moreover, a causal effect was deemed significant if the IVW p-value fell below the Bonferroni-corrected threshold (*p*□< □0.05/8□= □0.00625), coupled with consistent directionality in the weighted median and MR-Egger results.

#### Sensitivity analyses

We conducted sensitivity analyses to address horizontal pleiotropy and heterogeneity using weighted median, MR-Egger, and MR-PRESSO methods. These approaches help verify assumptions and assess robustness by identifying potential horizontal pleiotropy. The weighted median model provides consistent estimates when over half of the weights come from valid instrumental variables (IVs). MR-Egger regression detects and corrects horizontal pleiotropy, with its intercept term indicating directional pleiotropy if *p* < 0.05^20^. MR-PRESSO identifies and corrects outliers, offering outlier-corrected estimates and using a distortion test to compare estimation differences before and after outlier removal. Cochran’s Q test evaluates SNP heterogeneity for exposure, confirming consistency with MR assumptions if *p* < 0.05^16^. Consistent directionality across the three methods and the absence of horizontal pleiotropic effects are essential for credible causal inference.

## Results

### Genetic instrumental variable selection

Following the parameters described above, a total of 32 variants were used as IVs for various general anaesthetics. For spinal anaesthesia, 2 variants were used, and for local anaesthesia, 8 variants were employed(Fig. 2). All F-statistic values for the IVs were greater than 10, indicating no significant weak IV bias. Detailed information on these genetic variants used in the MR analyses is provided in Supplementary Tables 3, 5 and 7.

**Figure 2:**
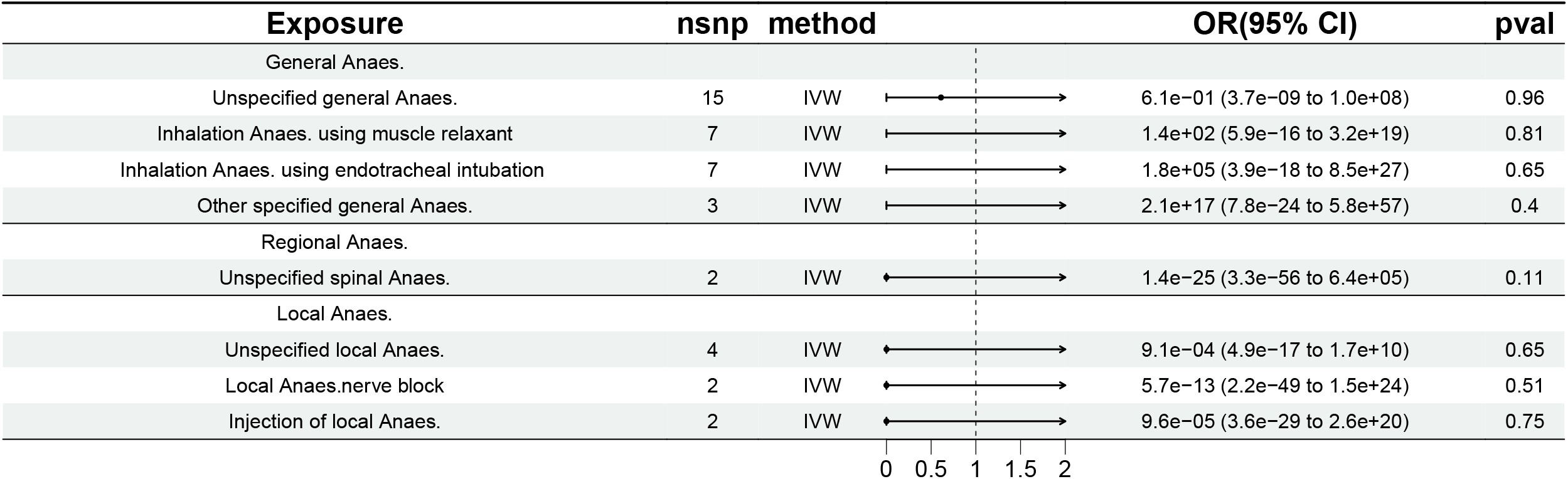
The causal effects of different anaesthesia types on delirium risk, with no significant associations observed. Anaes.:Anaesthesia, ; IVW: Inverse variance weighted; IVs: Instrumental variables; SNPs: Single Nucleotide Polymorphisms; MR, Mendelian randomization; TSMR, Two-Sample Mendelian randomization; OR: Odds ratio; CI, confidence interval; GWAS: Genome-Wide Association Study; UKBB, United Kingdom Biobank.

### Effects of genetically proxied anaesthesia on delirium

To explore the relationship between different anaesthesia subgroups and delirium, we analyzed data from the comprehensive UKBB database, which covers various types of anaesthesia. Our analysis found no causal effect of general, regional, or local anaesthesia on delirium (Fig. 2; Supplementary Tables 2, 4 and 6).

### Sensitivity analyses for MR estimates

Sensitivity analyses revealed consistent results with no evidence of horizontal pleiotropy, as indicated by the statistically insignificant Egger regression intercepts (Table 1). Heterogeneity was noted for the association between inhalation anaesthesia using endotracheal intubation and delirium (I^2^=60.9; Table 1), but results remained unchanged after excluding MR-PRESSO-identified outliers. Additionally, no heterogeneity was observed between other forms of anaesthesia and delirium, and leave-one-out tests showed no potential causality after excluding any SNP (Supplementary Figures 1-5).

## Discussion

Our MR study revealed no evidence of causal effects of anaesthesia on delirium risk, this conclusion was supported by the absence of genetic correlation between different forms of anaesthesia and delirium in our analysis. Importantly, our study marks the first documentation of the lack of a causal effect of anaesthesia on delirium.

The revelation that anaesthesia lacks a causal association with delirium challenges conventional wisdom and contradicts previous research findings^9,11,12,17^. Traditionally, different forms of anaesthesia have been linked to an increased risk of delirium, with patients undergoing deeper or longer anaesthesia facing a heightened risk from delirium compared to the general population. These observations have spurred a series of observational studies investigating the associations between anaesthesia and delirium, each offering valuable insights. For instance, a multicentre randomised clinical trial conducted in a hospital setting revealed a higher incidence of delirium among patients who received deep anaesthesia compared to those who underwent light anaesthesia anaesthesia^11^. Similarly, a cohort study among elderly patients subjected to general anaesthesia displayed more frequent cognitive impairment during the immediate postoperative period in comparison to those who received a regional technique^21^. These observational findings provide compelling evidence warranting further investigation into the role of anaesthesia in delirium onset and progression.

Our Mendelian randomization analysis challenges the presumed link between anaesthesia and delirium, prompting a reassessment. One possible explanation is the intricate interplay between anaesthesia and delirium. While anaesthesia is linked to certain surgical procedures like hip surgeries, it’s plausible that the surgery itself, rather than the anaesthesia, leads to delirium^22^. Additionally, differences in study design and biases between observational studies and Mendelian randomization could contribute to this discrepancy. Observational studies may be influenced by confounding factors such as age, commodities, and postoperative complications, whereas Mendelian randomization offers more dependable causal estimates by employing genetic variants as instrumental variables.

It’s essential to acknowledge that the absence of a causal effect doesn’t negate the potential risks associated with anaesthesia. Instead, it emphasizes the necessity for further investigation into the intricate relationship between anaesthesia, patient characteristics, and delirium development. Our study underscores the value of MR analysis in untangling potential causal pathways and providing a robust framework for comprehending the anaesthesia-delirium connection. Looking ahead, exploring specific anaesthetic agents, dosages, and administration protocols in relation to delirium risk could be beneficial. Investigating potential protective or mitigating factors influencing this relationship is also warranted. Additionally, future research should consider incorporating longitudinal data and diverse patient populations to enhance generalizability and provide a comprehensive understanding of factors contributing to delirium post-anaesthesia.

Our study acknowledges several limitations. Firstly, its focus on a single ethnic group may limit broader applicability, emphasizing the importance of replication in diverse cohorts. Secondly, the use of summary statistics from GWAS alone may introduce misclassification bias. Thirdly, external validation was not feasible due to constraints in available data. Lastly, concerns regarding potential pleiotropy in Mendelian randomization analysis and residual confounding persist, despite sensitivity analyses.

**In conclusion**, our two-sample MR analysis provides compelling evidence of no causal relationship between genetically determined anaesthesia and heightened risk of delirium, contrasting with most observational studies. To verify the accuracy of our results, future research using higher quality GWAS data and more advanced methods is necessary.

## Data Availability

All data produced in the present work are contained in the manuscript

https://gwas.mrcieu.ac.uk/

## Acknowledgments

We want to acknowledge the participants and investigators of the FinnGen study, UK Biobank, and other GWASs included in this study.

## Data Availability Statement

All data generated or analyzed during this study are included in this article and its supplementary information files.

## Ethics Statement

Not applicable

## Author Contributions

L.T. conceived the project, designed the study and revised the manuscript.; YH.L. wrote the manuscript; CY.Z. collected data from the public database; YW.S., and YH.L. performed the study, and all authors reviewed the manuscript.

## Consent for Publication

Not applicable

### Abbreviations

anaesthesia: anaes.
IVW: Inverse variance weighted
IVs: Instrumental variables
SNPs: Single Nucleotide Polymorphisms
MR: Mendelian randomization
TSMR: Two-Sample Mendelian randomization
OR: Odds ratio
CI: confidence interval
GWAS: Genome-Wide Association Study
UKBB: United Kingdom Biobank

