## Supplementary Tables and figures for "Anaesthesia and delirium: a Mendelian randomization study": Supplementary Figures.pdf

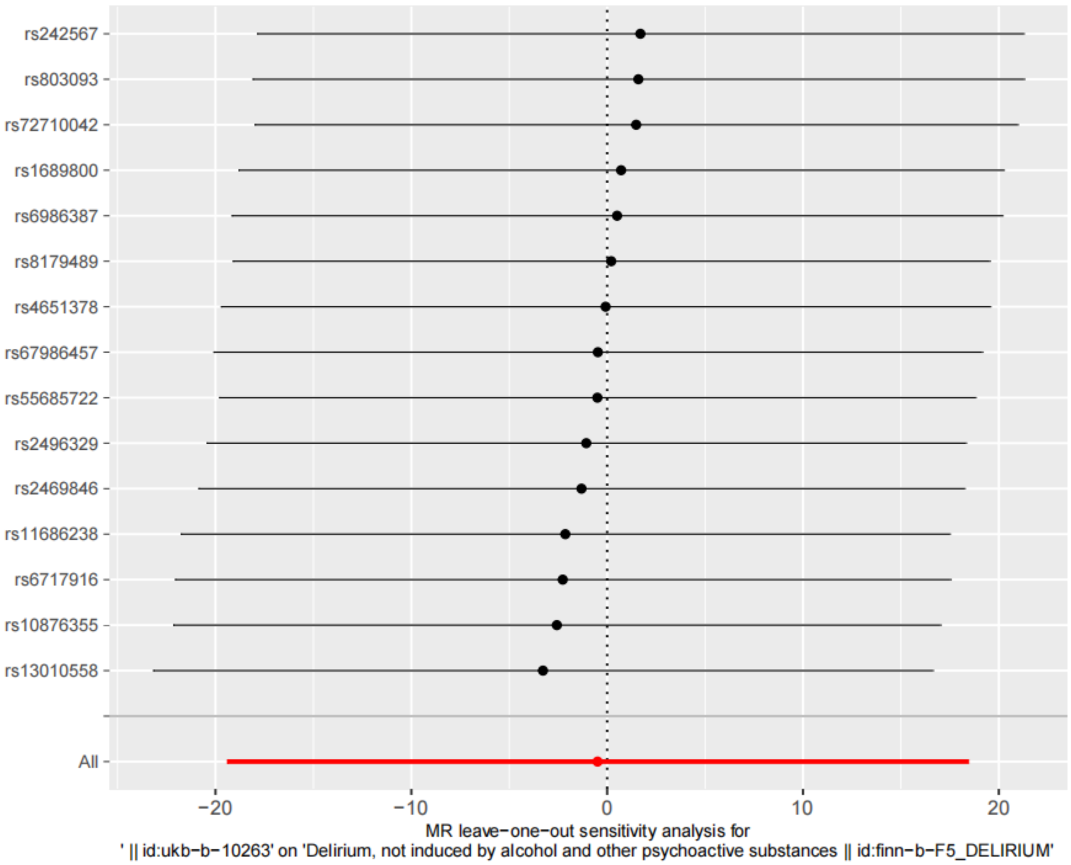


Supplementary Figures 1. Leave-one-out sensitivity analysis plots for Unspecified general Anaes. on delirium. Anaes.: Anaesthesia; MR, mendelian randomization.

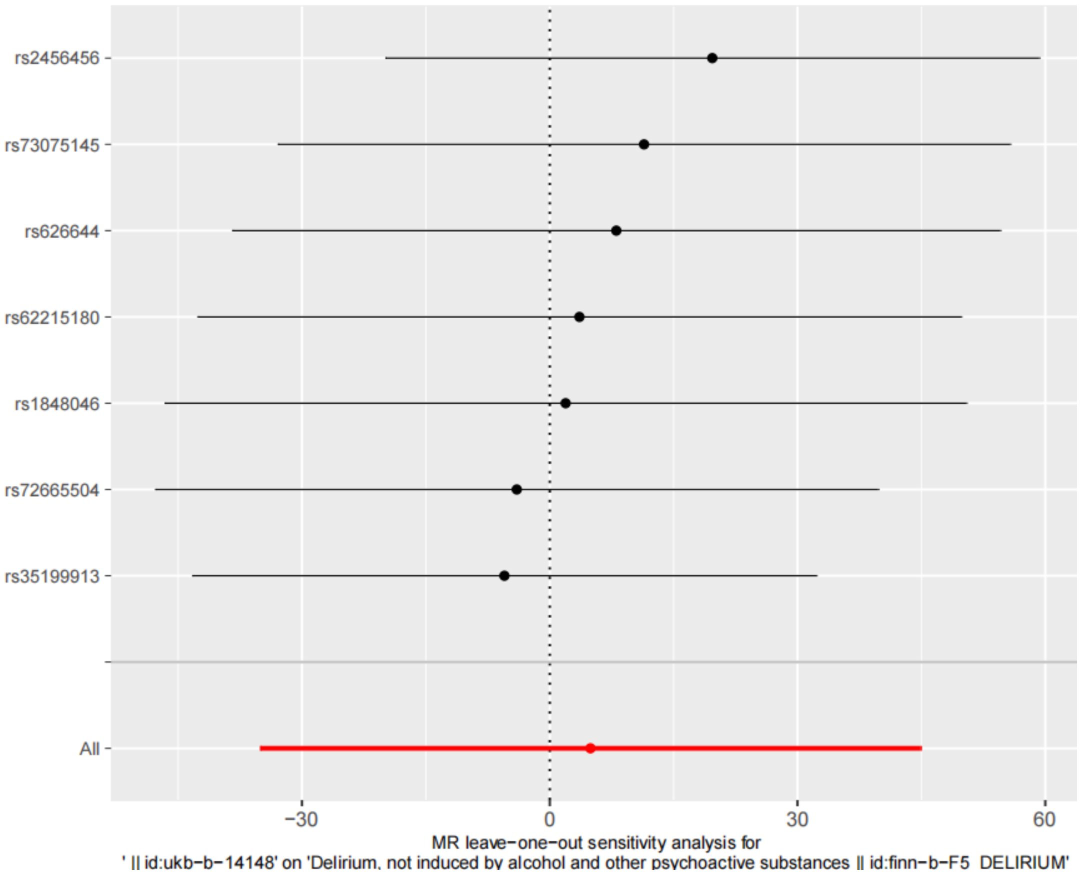


Supplementary Figures 2. Leave-one-out sensitivity analysis plots for Inhalation Anaes. using muscle relaxant on delirium. Anaes.: Anaesthesia; MR, mendelian randomization.

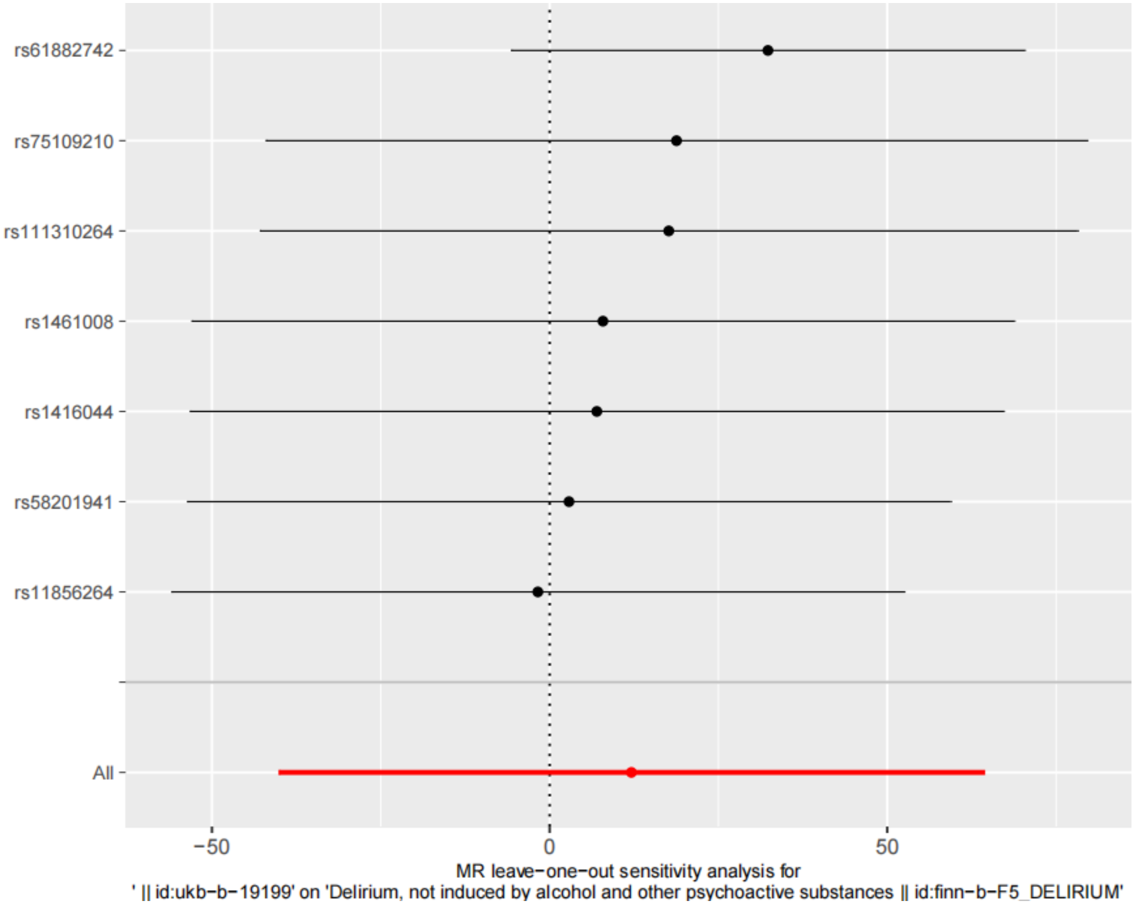


Supplementary Figures 3. Leave-one-out sensitivity analysis plots for Inhalation Anaes. using endotracheal intubation on delirium. Anaes.: Anaesthesia; MR, mendelian randomization.

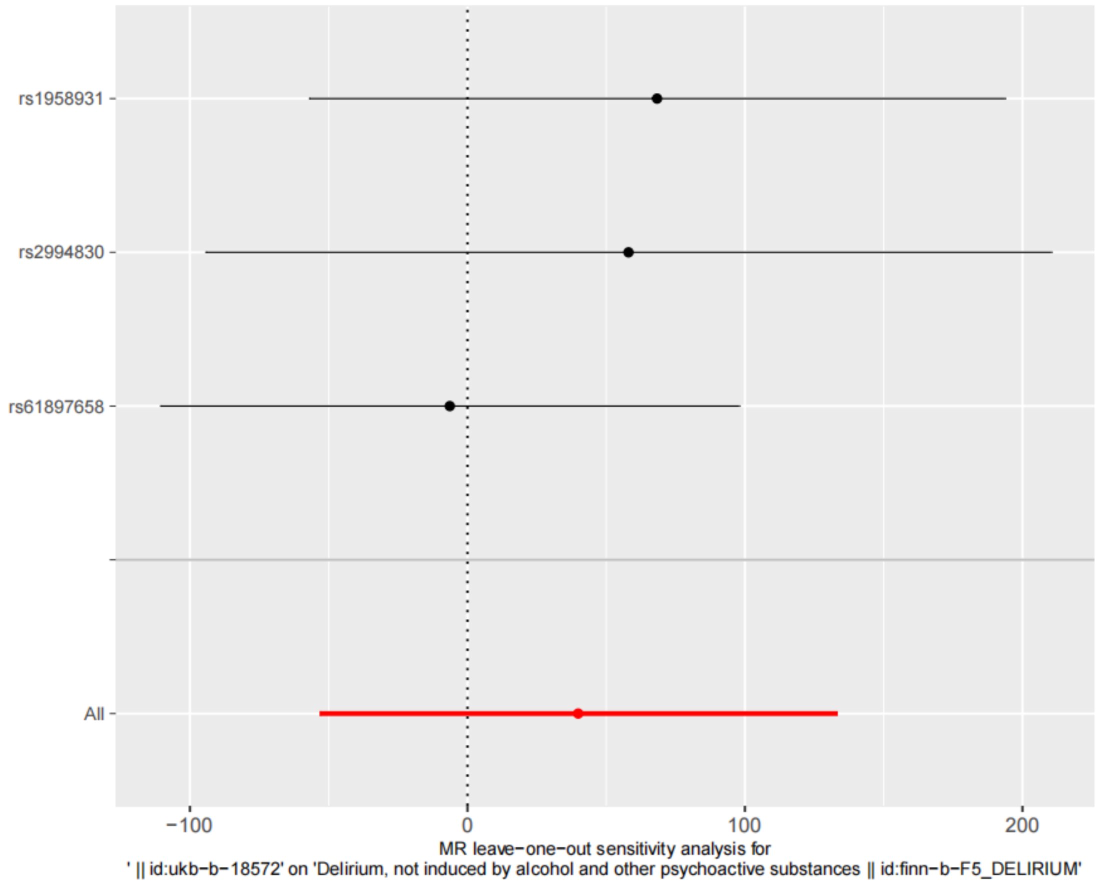


Supplementary Figures 4. Leave-one-out sensitivity analysis plots for Other specified general Anaes. on delirium. Anaes.: Anaesthesia; MR, mendelian randomization.

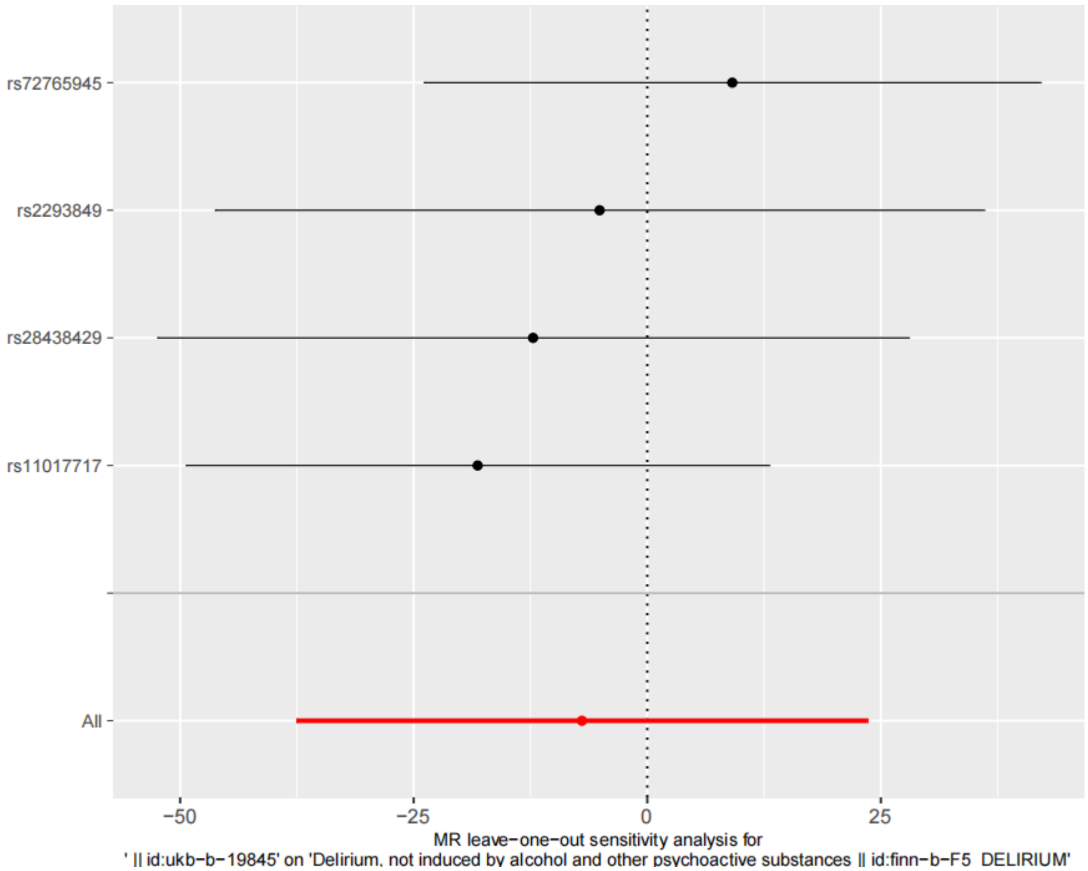

Supplementary Figures 5. Leave-one-out sensitivity analysis plots for Unspecified local Anaes. on delirium. Anaes.: Anaesthesia; MR, mendelian randomization.
